# Inflammatory Markers are Associated with Ischemic Stroke among Indigenous Africans: Evidence from the SIREN Study

**DOI:** 10.1101/2025.08.10.25331295

**Authors:** Moustafa I. Morsy, Fred S. Sarfo, Osahon J. Asowata, Onoja Akpa, Joshua Odunayo Akinyemi, Albert Akpalu, Olayinka Adebajo, Oladimeji Adebayo, Akinkunmi Okekunle, Kazeem Akinwande, Paul Olowoyo, Ganiyu A Amusa, Reginald Obiako, Oyedunni S. Arulogun, Daniel T. Lackland, Isah S. Yahaya, Abiodun M. Adeoye, Olayemi Balogun, Elisabetta Caiazzo, Lucy McShane, Philip Ibinaiye, Benedict Calys-Tagoe, Yaw Mensah, Adekunle G. Fakunle, Mayowa Ogunronbi, Osi Adeleye, Samuel Diala, Joseph Yaria, Oladotun Olalusi, Akintomiwa Makanjuola, Godwin Ogbole, Michael B. Fawale, Ayomide M. Owolabi, Lukman Owolabi, Okechukwu S. Ogah, Atinuke Agunloye, Morenikeji Komolafe, Innocent Chukwuonye, Godwin Osaigbovo, Giuseppe D’Agostino, Pierpaolo Pellicori, Dario Bruzzese, Rufus Akinyemi, Tomasz J. Guzik, Hemant K. Tiwari, Bruce Ovbiagele, Pasquale Maffia, Mayowa O. Owolabi

## Abstract

**Background:** Inflammation might predispose to worse outcomes after an ischemic stroke. This has not been characterized among indigenous Africans.

**Purpose:** We investigated the association between inflammatory biomarkers, stroke severity, and outcomes in Africans.

**Methods:** This is a retrospective analysis of a prospective study, including 90 participants with confirmed ischemic stroke selected from the Stroke Investigative Research & Educational Network (SIREN) cohort and 90 controls matched by age, sex, and ethnicity. Plasma concentrations of 368 protein biomarkers were analyzed (Olink® Explore 384 Inflammation panel) and compared between cases and controls. Further, we investigated the association between these proteins and stroke severity, lesion volume, and one-month post-stroke disability and fatality.

**Results:** Differential protein expression analysis revealed 23 up-regulated and 14 down-regulated proteins in stroke cases versus controls. Among the up-regulated proteins, agouti-related protein (AgRP) and tumor necrosis factor receptor superfamily member 11A (TNFRSF11A) were the most significantly up-regulated, while interleukin-1 beta (IL-1β) and integrin alpha-11 (ITGA11) were the most down-regulated proteins. Logistic regression analysis identified 72 proteins that were associated with stroke severity independent of age and sex. Among these, complement C1q subcomponent subunit A (C1qa) exhibited the strongest associations. Additionally, 14 proteins including C-C motif chemokine 23 (CCL23) were found to be associated with one-month post-stroke disability.

**Conclusion:** Stroke disability and severity were associated with inflammation in an indigenous African population. The identified biomarkers might be useful for predicting stroke outcome or serve as therapeutic target.

**Graphical abstract:** 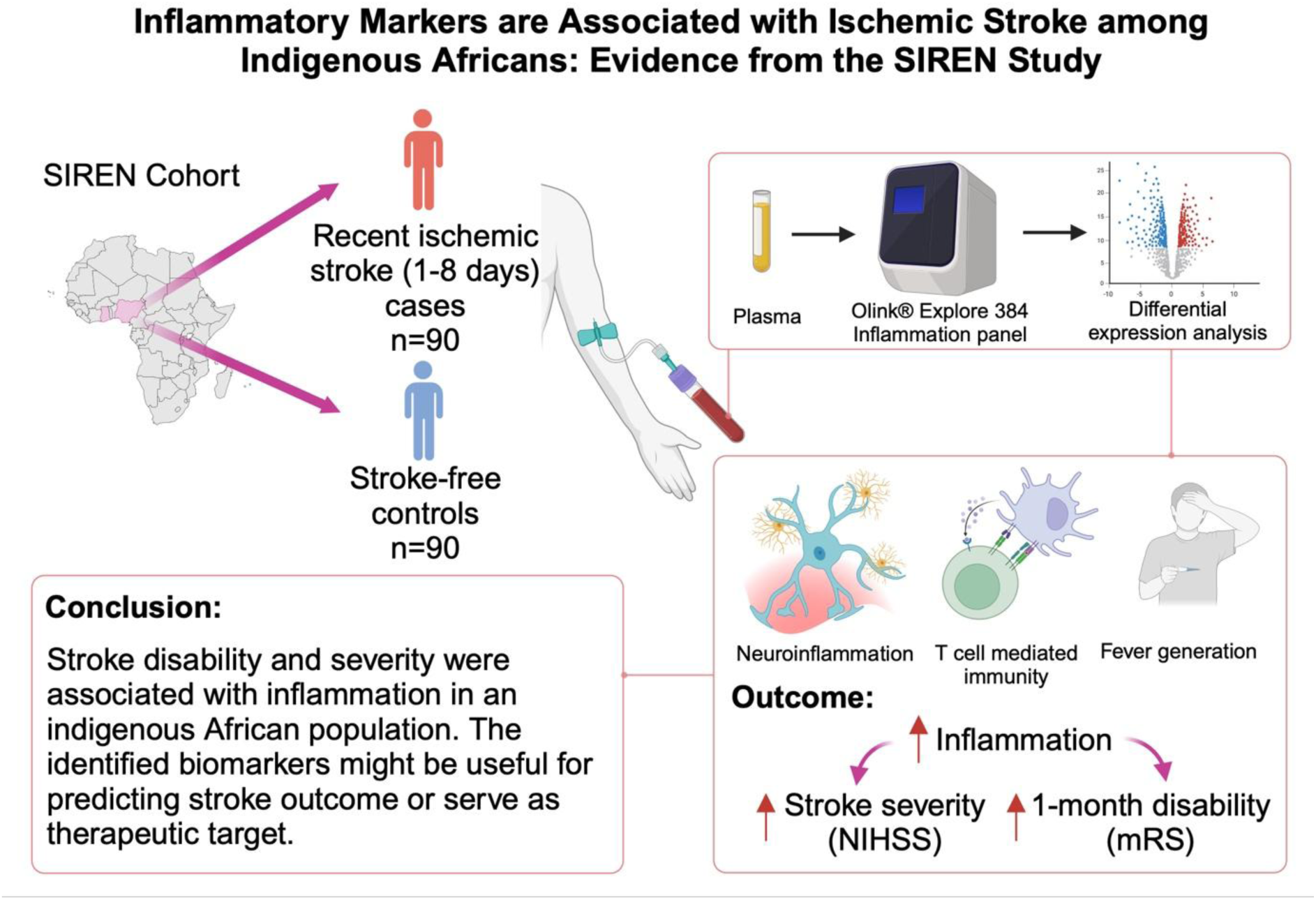

## INTRODUCTION

Stroke is a major cause of death, dementia, and disability worldwide.^1^ Inflammation is pivotal to the pathophysiology and outcome of ischemic stroke, the more common stroke phenotype.^2^ Inflammatory cells orchestrate the elaboration of several inflammatory proteins that are involved in arterial occlusion, cerebral parenchymal injury, subsequent tissue repair, and post-stroke complications including depression and dementia.^3^

Africa experiences one of the highest global burdens of stroke, with onset occurring at a younger age and mortality rates at least five times greater than those in high-income countries.^1^ The region’s epidemiological profile reflects a dual burden of communicable and non-communicable diseases, with chronic inflammation potentially serving as a common pathway that predisposes individuals to increased stroke occurrence and poor outcomes.^4–6^ Contributing factors include an elevated risk of cardiometabolic disorders driven by infections, malnutrition, sedentary lifestyles, smoking, and air pollution, which are particularly prevalent in Africa and further exacerbate stroke risk through the promotion of endothelial dysfunction and inflammation.^7–9^ Emerging lines of evidence posit polymorphisms in genes coding for IL-6 and IL-1β in Africans to be associated with stroke.^10,11^ Elucidating the profile of inflammatory markers associated with severity of stroke and its outcomes among indigenous Africans may provide useful mechanistic insights and aid with potential biomarker discovery in this population. In the present study, we aimed to characterize the circulating inflammatory biomarkers following the onset of stroke and their associations with stroke outcomes.

## METHODS

### Study population and study design

The Stroke Investigative Research and Educational Networks (SIREN) study is a prospective case-control study that included patients hospitalized with acute ischemic or hemorrhagic stroke in 16 collaborating sites in Nigeria and Ghana. Out of 1430 cases with confirmed ischemic stroke, a subset of 90 cases with accessible genome data, and matched stroke-free controls were selected for the present analysis. The study protocol has been published elsewhere.^12^ Briefly, consent was obtained from consecutive stroke cases for the study including unconscious and aphasic patients whose next-of-kin consented. All cases were adults aged ≥18 years with clinical stroke presenting within 8 days of current symptom onset or ‘last seen without deficit’. All stroke diagnoses were confirmed using computed tomography (CT) or magnetic resonance imaging (MRI) scans, typically within 10 days of symptom onset. Controls were consenting stroke-free adults, mainly from the communities in the catchment areas of the hospitals where cases were recruited. Hospital-based controls were either attendants or relatives of another (non-stroke) patient or patients admitted to the hospital or visiting the hospital for conditions or procedures not related to stroke or transient ischemic stroke (TIA). Stroke-free status was confirmed with the 8-item questionnaire for verifying stroke-free status (QVSFS) which has a 98% negative predictive value.^13^ Exclusion criteria for controls included current hospitalization for coronary heart disease or known previous history of stroke. Controls were matched to cases by age (+/- 5 years), sex, and ethnicity in a ratio of 1:1.

For baseline risk factors, diabetes mellitus (DM) was defined by a history of DM, use of DM medications, HBA1c >7%, or fasting blood glucose (FBG) ≥7.0mmol/L. FBG was measured after 7 days after stroke to account for stress-induced transient glucose rise. Dyslipidemia was defined as fasting total cholesterol ≥5.2mmol/L, low-density lipoprotein ≥3.4mmol/L, high-density lipoprotein ≤1.03mmol/L, triglycerides ≥1.7mmol/L (as per the National Cholesterol Education Program guidelines^14^) or use of statin prior to stroke onset. Hypertension was defined using a cutoff of ≥140/90 mmHg 72 hours after stroke, a history of hypertension, or use of antihypertensive drugs before stroke or >72 hours after stroke. Cardiac disease was defined based on either a history, clinical examination, and electrocardiogram (ECG) or echocardiographic evidence of cardiac arrhythmias (mainly atrial fibrillation or flutter), heart failure, cardiomyopathy, or valvular heart diseases. Smoking and alcohol former or current use were defined as self-reported.

Stroke severity was assessed using the National Institute of Health Stroke Scale (NIHSS)^15^ and the Stroke Levity Scale (SLS).^16^ A stroke case was considered severe if the NIHSS score was >15 or the SLS was ≤5. Subtypes of ischemic stroke were classified according to Trial of ORG 10172 in Acute Stroke Treatment (TOAST).^17^ Neurologic disability 1 month after stroke onset was assessed using the modified Rankin Scale (mRS).^18^ Lesion volume was calculated using the ellipsoid equation and classified as small (≤10 cm^3^), medium(10.1-30 cm^3^), or large(>30 cm^3^).^19^

Ethical approval was obtained from all study sites and informed consent was obtained from all participants before enrolment. The study was approved by the University of Ibadan and University College Hospital Ibadan, Nigeria.

### Protein biomarkers analysis

For proteomics analysis, plasma samples were collected within 1-8 days of stroke onset, stored at - 80°C, and transferred to Olink Bioscience analysis service (Uppsala, Sweden). Samples were analyzed using Olink® Explore 384 Inflammation panel, which contains a panel of 368 unique protein biomarkers (Table S1). Olink multiplex assay is based on proximity extension assay (PEA) technology that links protein-specific antibodies to DNA-encoded tags which bypasses the limitation of cross-reactivity in conventional multiplexed immunoassays ensuring no loss in specificity and sensitivity of the assay.^20^ The assay quantifies protein expression in normalized protein expression (NPX) to account for intra and inter-assay variations. Since NPX is measured in log2 scale, 1 unit change in NPX difference equals a doubling of protein concentration. The full list of biomarkers assessed is in Table S1. Assays were run blinded from case and control status. For the 368 proteins measured, 2 proteins that were below the limit of detection (LOD) were excluded from the analysis. The LOD is defined by the 3 negative controls run on each plate and set to 3 SDs above the measured background. In addition, proteins with values <LOD in >50% of the samples in cases and controls were excluded (n=30). Principal component analysis (PCA) was employed for outlier detection where samples laying ≥3 standard deviations (SD) from the mean of PC1 and PC2 were excluded (n=1).

### Statistical analysis

Analysis was performed using R statistical software version 4.4.2. Baseline characteristics were reported as mean and standard deviation (SD) or median and interquartile range (IQR) for continuous variables; or number (percentage) for categorical variables. For between-group comparisons, the t-test or Mann-Whitney test was used for continuous variables while χ2 or Fischer exact tests for categorical. Differential expression analyses were conducted using Linear models for microarray analysis software R package (Limma version 3.60.4). The analysis was corrected for multiple testing using the Benjamini-Hochberg method. Proteins with a false discovery rate (FDR) <0.05 and minimum fold change (FC) of 1.4 were considered significant. Correlation between differentially expressed proteins (DEP) and population characteristics was performed using Spearman’s rank correlation. In case of missing values, no data imputation was performed. Tests were 2-sided, and *P* values <0.05 were considered significant.

#### Pathway overrepresentation analyses

To investigate the overrepresented pathway associated with DEP between cases and controls, the query of Gene Ontology (GO) biological processes was performed using (ClueGO version 2.5.10) plugin and (Cytoscape version 3.10.2) software. Fischer exact test correcting for multiple comparisons was used to find significantly enriched pathways. Pathways from “all experiments” and with FDR ≤ 0.01 were chosen.

#### Outcome analysis

Binary and ordinal logistic regression models were used to address the association between inflammatory biomarkers with stroke severity, disability at 1-month post-stroke, lesion volume and mortality, as appropriate. Multivariable models were adjusted for age and sex.

## RESULTS

In this analysis, 89 stroke cases and 90 stroke-free controls were included. The mean (SD) age of cases was 59.4 (13.0) years and 58.1 (13.6) years for controls with 48% of cases being females and 49% females among controls. Cases differed from controls in the location of residence, family history of stroke, and vascular risk factors, including diabetes mellitus, hypertension, and dyslipidemia, as shown in Table 1.

**Table 1.** Baseline characteristics of the included cohort.

| **Characteristic** | **Available (N)** | **Case, N = 89** | **Control, N = 90** | **p-value** |
| --- | --- | --- | --- | --- |
| Country (Nigeria), n (%) | 179 | 85 (96) | 86 (96) | >0.9 |
| Age, years, mean (SD) | 179 | 59 (13) | 58 (14) | 0.5 |
| Women, n (%) | 179 | 43 (48) | 44 (49) | 0.9 |
| Domicile, n (%) | 179 |  |  | **0.007** |
| Rural |  | 10 (11) | 27 (30) |  |
| Semi-urban |  | 30 (33) | 27 (30) |  |
| Urban |  | 50 (56) | 36 (40) |  |
| Smoking, n (%) | 179 | 10 (11) | 6 (7) | 0.3 |
| Alcohol use, n (%) | 178 | 21 (24) | 21 (24) | >0.9 |
| Family history of stroke, n (%) | 179 | 20 (22) | 8 (9) | **0.012** |
| Family history of CVD, n (%) | 179 | 30 (34) | 27 (30) | 0.6 |
| Dyslipidemia, n (%) | 179 | 66 (74) | 40 (44) | **<0.001** |
| DM, n (%) | 179 | 38 (43) | 3 (3) | **<0.001** |
| Hypertension, n (%) | 179 | 82 (92) | 66 (73) | **<0.001** |
| CVD, n (%) | 178 | 4 (5) | 6 (7) | 0.7 |
| BMI, kg/m<sup>2</sup>, mean (SD) | 175 | 26.8 (5.0) | 26.23 (6.0) | 0.5 |
| SBP, mmHg, median (IQR) | 179 | 150<br>(130-160) | 140<br>(120-160) | **0.029** |
| DBP, mmHg, median (IQR) | 179 | 90<br>(80-100) | 85<br>(79-93) | **0.01** |
| Total cholesterol, mmol/L, mean (SD) | 147 | 4.6 (1.1) | 4.6 (1.0) | 0.9 |
| Triglyceride, mmol/L, median (IQR) | 149 | 1.2 (0.9-1.5) | 1.2 (0.8-1.4) | 0.3 |
| HDL, mmol/L, mean (SD) | 149 | 1.2 (0.4) | 1.4 (0.5) | **0.014** |
| LDL, mmol/L, mean (SD) | 142 | 2.9 (1.0) | 2.8 (0.9) | 0.4 |
| Antidiabetic, n (%) | 178 | 10 (11) | 1 (1) | **0.005** |
| Antihyperlipidemic, n (%) | 179 | 2 (2) | 0 (0) | 0.2 |
| Stroke subtypes (TOAST), n (%) | 89 |  |  | - |
| Large-artery atherosclerosis |  | 43 (48) | - |  |
| Small-artery occlusion |  | 46 (52) | - |  |

BMI, Body mass index; CVD, cardiovascular disease; DBP, diastolic blood pressure; DM, diabetes mellites; HDL, high-density lipoprotein; LDL, low-density lipoprotein; SBP, systolic blood pressure; DBP, diastolic blood pressure, TOAST, Trial of ORG 10172 in Acute Stroke Treatment.

### Profile of inflammatory protein expression between cases and controls

The protein profiles of stroke cases and stroke-free controls shows partial separation from principal component analysis (Figure 1A). In differential expression analysis, stroke cases showed 23/336 up-regulated proteins and 14/336 down-regulated compared to stroke-free controls (Figure 1B). Agouti-related peptide (AGRP) (logFC 0.94) and TNF receptor superfamily member 11a (TNFRSF11A) (logFC 0.84) were the most up-regulated proteins, while IL-1β (logFC -1.37) and Integrin α_11_ (ITGA11) (logFC -1.15) were the most down-regulated (Figure 1C and Table S2). Correlation matrix between differentially expressed proteins and subjects’ clinical characteristics are provided in (Figure S1 & Table S3). Notably, ITGA11 positively correlated with HDL in stroke cases (r= 0.40), while marginal zone B and B1 cell specific protein (MZB1) correlated negatively (r= -0.46).

**Figure 1.**
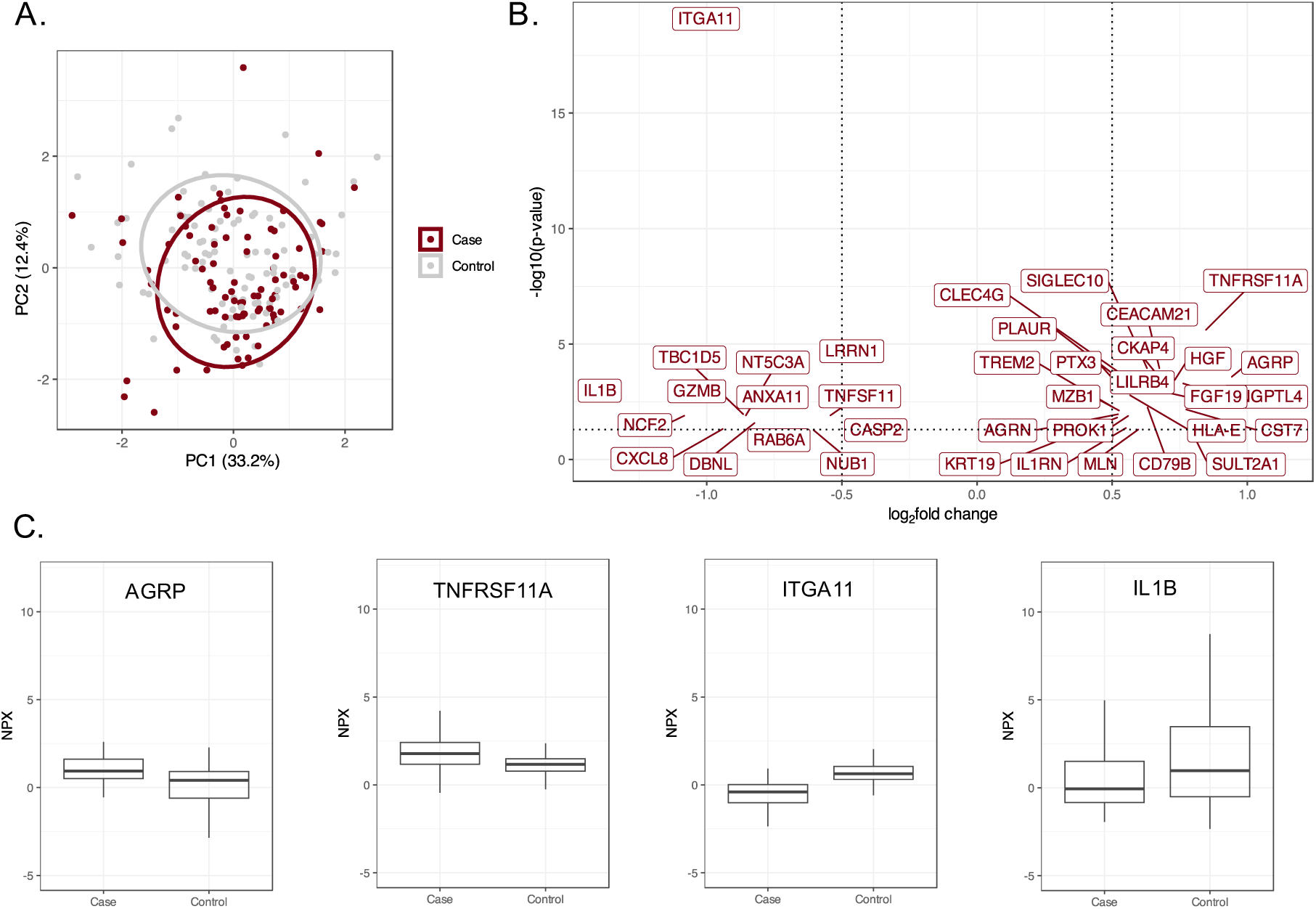
Differentially expressed inflammatory proteins between stroke cases and controls. (A) Principal component analysis of plasma inflammatory proteins using the first 2 principal components. (B) Volcano plot of differentially expressed inflammatory proteins between stroke cases and controls. Proteins above log2fold change = 0.5 (fold change 1.4) and false discovery rate (FDR) < 0.05 were annotated. (C) Boxplots of Normalized Protein eXpression (NPX) of the most upregulated and downregulated proteins in stroke cases and controls.

In pathway representation analysis of differentially expressed proteins between cases and controls, a total of 35 pathways were identified (Table S4). The strongest pathways associated with stroke were the regulation of T cell-mediated immunity, positive regulation of fever generation, and regulation of neuroinflammatory response (Figure 2). The majority of the pathways were related to IL-β, TNF Receptor superfamily member 11α (TNFRSF11A), and its ligand TNF superfamily member 11 (TNFSF11).

**Figure 2.**
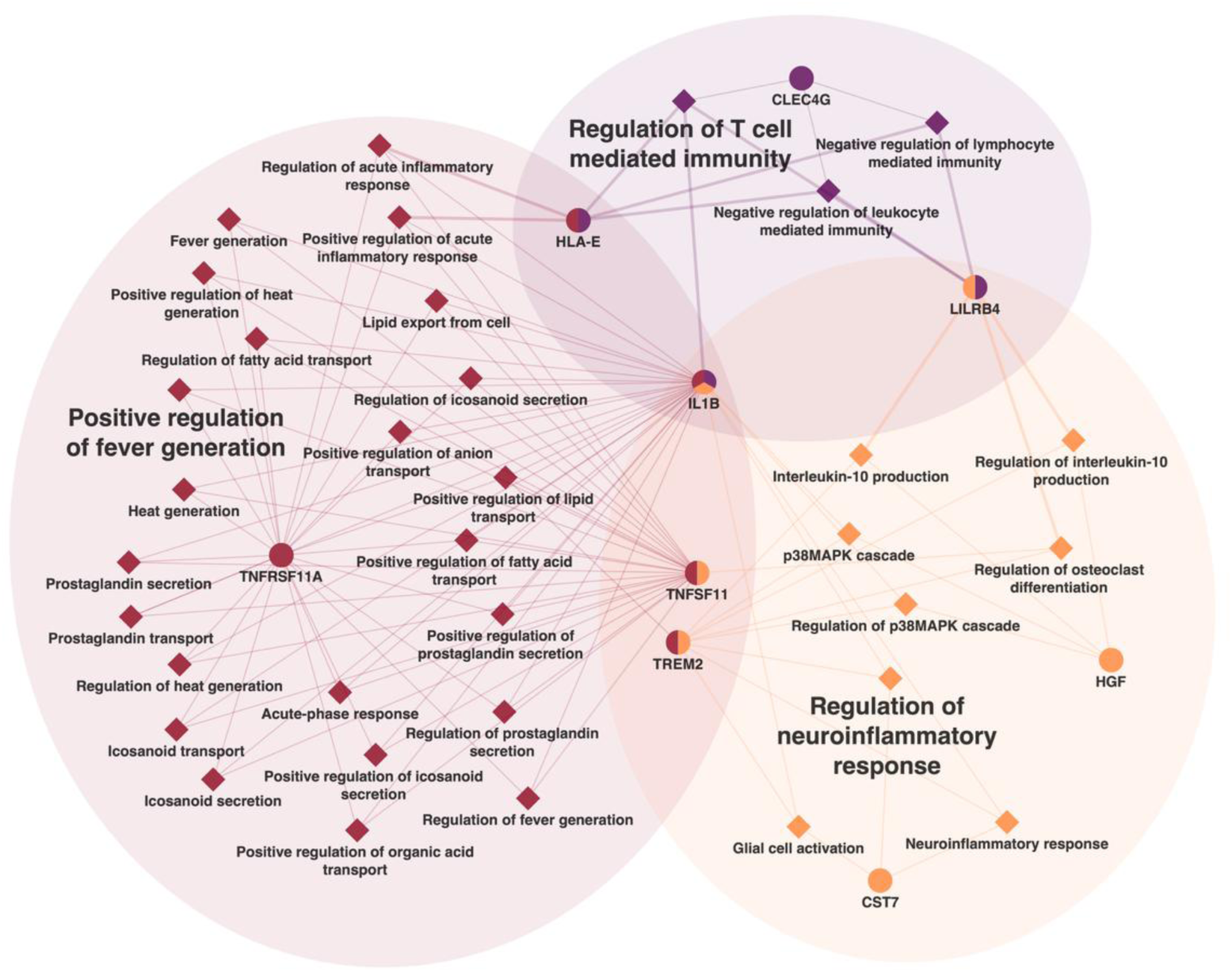
Pathway enrichment analysis of the most upregulated and downregulated inflammatory proteins in stroke cases and controls, and the major proteins connecting the pathways.

### Association of inflammatory protein biomarkers with stroke severity

Out of the 89 stroke cases analyzed, 19 (21.4%) exhibited a severe form of stroke as assessed by the NIHSS scale, while 28 (31.5%) were classified as severe based on the SLS scale (Table 2 & Table S5). Notably, 9 (10.1%) cases had missing NIHSS scores, and 4 (4.5%) cases had missing SLS scores. Severe stroke cases had lower levels of total cholesterol and low-density lipoprotein (LDL) compared to non-severe cases.

**Table 2:** Baseline characteristics of cases by stroke severity on National Institutes of Health Stroke Scale (NIHSS)

|  | NIHSS |  |  |
| --- | --- | --- | --- |
| Characteristic | Severe, N = 19 | Non-severe, N = 61 | p-value |
| Age, years, mean (SD) | 63 (14) | 58 (13) | 0.2 |
| Women, n (%) | 8 (42) | 28 (46) | 0.8 |
| Domicile, n (%) |  |  | 0.8 |
| Rural | 1 (5) | 8 (13) |  |
| Semi-urban | 7 (37) | 20 (33) |  |
| Urban | 11 (58) | 33 (54) |  |
| Smoking, n (%) | 2 (11) | 8 (13) | >0.9 |
| Alcohol use, n (%) | 5 (26) | 15 (25) | >0.9 |
| Family history of stroke, n (%) | 5 (26) | 12 (20) | 0.5 |
| Family history of CVD, n (%) | 6 (32) | 20 (33) | >0.9 |
| Dyslipidemia, n (%) | 14 (74) | 45 (74) | >0.9 |
| DM, n (%) | 10 (53) | 23 (38) | 0.2 |
| Hypertension, n (%) | 17 (89) | 56 (92) | 0.7 |
| Cardiac disease, n (%) | 2 (11) | 2 (3) | 0.2 |
| BMI, kg/m<sup>2</sup>, mean (SD) | 25.6 (5.4) | 27.6 (4.9) | 0.2 |
| SBP, mmHg, median (IQR) | 157 (145-174) | 147 (130-160) | 0.12 |
| DBP, mmHg, median (IQR) | 97 (83-100) | 90 (80-100) | 0.2 |
| Total cholesterol, mmol/L, mean (SD) | 3.9 (1.1) | 4.8 (1.1) | **0.009** |
| Triglyceride, mmol/L, median (IQR) | 1.1 (1.0-1.7) | 1.2 (0.9-1.5) | 0.6 |
| HDL, mmol/L, mean (SD) | 1.2 (0.5) | 1.2 (0.4) | >0.9 |
| LDL, mmol/L, mean (SD) | 2.2 (0.9) | 3.0 (1.1) | **0.013** |
| Stroke subtypes (TOAST), n (%) |  |  | 0.2 |
| Large-artery atherosclerosis | 11 (58) | 26 (43) |  |
| Small-artery occlusion | 8 (42) | 35 (57) |  |
| Lesion volume, cm<sup>3</sup>, n (%) |  |  | 0.4 |
| <10 | 10 (67) | 42 (82) |  |
| 10-30 | 3 (20) | 5 (9) |  |
| >30 | 2 (13) | 4 (8) |  |

BMI, Body mass index; CVD, cardiovascular disease; DBP, diastolic blood pressure; DM, diabetes mellites; HDL, high-density lipoprotein; LDL, low-density lipoprotein; SBP, systolic blood pressure; TOAST, Trial of ORG 10172 in Acute Stroke Treatment.

**Table 3:** Multivariable OR and 95% CI for the biomarkers associated with severity of stroke according to National Institutes of Health Stroke Scale (NIHSS)

| Biomarker | NIHSS |  |
| --- | --- | --- |
|  | OR (95 CI%) | BH-adjusted P value |
| AGER | 2.61 (1.47-5.20) | 0.015 |
| AGRN | 2.4 (1.47-4.42) | 0.015 |
| AGRP | 2.2 (1.39-3.94) | 0.015 |
| ANGPTL4 | 3.94 (1.98-9.52) | 0.012 |
| B4GALT1 | 4.97 (2.18-14.22) | 0.012 |
| C1QA | 50.76 (6.95-616.65) | <0.001 |
| CCL23 | 4.26 (2.10-10.77) | <0.001 |
| CCL7 | 2.51 (1.51-4.63) | 0.012 |
| CD40 | 3.5 (1.82-8.00) | 0.012 |
| CD70 | 3.64 (1.66-9.18) | 0.021 |
| CD79B | 1.83 (1.23-2.92) | 0.028 |
| CDSN | 2.12 (1.29-3.76) | 0.028 |
| CEACAM21 | 2.07 (1.23-3.82) | 0.047 |
| CKAP4 | 3.34 (1.85-7.23) | <0.001 |
| CLEC4C | 0.34 (0.17-0.62) | 0.012 |
| CLEC4G | 8.18 (3.28-27.15) | <0.001 |
| COLEC12 | 3.69 (1.69-9.14) | 0.015 |
| CRELD2 | 2.61 (1.53-4.96) | 0.012 |
| CRIM1 | 4.3 (1.86-11.87) | 0.015 |
| CSF1 | 4.72 (1.99-13.36) | 0.012 |
| CXADR | 2.32 (1.38-4.22) | 0.021 |
| ENAH | 3.28 (1.80-6.92) | <0.001 |
| ESM1 | 2.61 (1.44-5.52) | 0.025 |
| F2R | 1.79 (1.19-2.84) | 0.041 |
| FABP1 | 1.74 (1.22-2.63) | 0.025 |
| FCAR | 3.86 (1.80-9.76) | 0.012 |
| FST | 2.36 (1.32-4.69) | 0.038 |
| FSTL3 | 3.73 (2.00-8.00) | <0.001 |
| HSD11B1 | 3.48 (1.57-8.81) | 0.025 |
| HSPA1A | 1.55 (1.12-2.19) | 0.047 |
| IFNGR1 | 4.66 (2.04-13.01) | 0.012 |
| IL10RB | 2.68 (1.34-5.92) | 0.041 |
| IL12RB1 | 2.79 (1.42-6.17) | 0.028 |
| IL15 | 5.69 (2.44-17.75) | 0.012 |
| IL17RB | 2.58 (1.49-5.09) | 0.015 |
| IL18R1 | 4.61 (1.63-14.97) | 0.033 |
| IL4R | 3.13 (1.48-7.34) | 0.028 |
| IL5RA | 2.32 (1.28-4.57) | 0.041 |
| IL6 | 2.02 (1.42-3.09) | <0.001 |
| ITGA11 | 0.3 (0.12-0.66) | 0.025 |
| JUN | 3.35 (1.88-7.11) | <0.001 |
| KRT19 | 2.06 (1.35-3.52) | 0.015 |
| LAIR1 | 1.9 (1.19-3.2) | 0.047 |
| LGALS9 | 3.69 (1.73-8.88) | 0.015 |
| LIFR | 5.22 (2.02-16.33) | 0.015 |
| LILRB4 | 3.86 (1.98-9.41) | 0.012 |
| LTBR | 3.4 (1.85-7.35) | <0.001 |
| LY6D | 2.14 (1.34-3.66) | 0.015 |
| MAPK9 | 2.18 (1.24-4.04) | 0.045 |
| MATN2 | 4.27 (1.99-11.87) | 0.012 |
| MERTK | 3.27 (1.68-7.44) | 0.015 |
| MILR1 | 2.24 (1.45-3.81) | 0.012 |
| NPPC | 1.59 (1.15-2.37) | 0.047 |
| NUDC | 2.11 (1.28-3.77) | 0.033 |
| PADI2 | 2 (1.29-3.35) | 0.025 |
| PGF | 3.37 (1.77-7.41) | 0.012 |
| PLAUR | 4.72 (2.23-12.73) | <0.001 |
| PRELP | 4.41 (1.87-12.69) | 0.015 |
| PROK1 | 2.67 (1.57-5.20) | 0.012 |
| PTX3 | 3.97 (1.80-10.71) | 0.015 |
| SCGB1A1 | 3.46 (1.78-7.97) | 0.012 |
| SIGLEC10 | 2.47 (1.44-4.66) | 0.015 |
| SPINT2 | 2.04 (1.32-3.39) | 0.021 |
| STX8 | 1.93 (1.24-3.17) | 0.028 |
| TLR3 | 0.23 (0.07-0.65) | 0.047 |
| TNFRSF11A | 2.98 (1.71-6.01) | 0.012 |
| TNFRSF11B | 3.86 (1.78-9.84) | 0.015 |
| TNFRSF14 | 2.94 (1.58-6.19) | 0.015 |
| TNFSF11 | 0.49 (0.3-0.76) | 0.015 |
| TNFSF13 | 2.67 (1.45-5.63) | 0.025 |
| VEGFA | 1.99 (1.22-3.46) | 0.045 |
| WNT9A | 2.45 (1.43-4.67) | 0.021 |

Models adjusted for age and sex. Biomarkers names are available in Table S1. NIHSS, National Institutes of Health Stroke Scale.

After adjusting for age and sex, 72 proteins were associated with stroke severity on the NIHSS scale. Complement C1q subcomponent subunit A (C1QA) and C-type lectin domain family 4 member G (CLEC4G) exhibited the strongest positive associations, while Toll-like receptor 3 (TLR3) and ITGA11 exhibited the strongest negative associations. Classification of severity on the SLS scale yielded only CLEC4G as a significant predictor of stroke severity (Table S6).

### Association of inflammatory protein biomarkers with ischemic lesion volume

Lesion volume was measured in 75 cases (83.33%). No proteins were identified as significant predictors of lesion size after controlling for false discovery rate (Table S7).

### Association of inflammatory protein biomarkers with neurologic disability at 1-month post stroke

Higher expression of c-c motif chemokine 23 (CCL23), plasminogen activator, urokinase receptor (PLAUR), matrilin-2 (MATN2), CLEC4G, Protein enabled homolog (ENAH), c-c motif chemokine 7 (CCL7), cytoskeleton-associated protein 4 (CKAP4), tumor necrosis factor receptor superfamily member 3 (LTBR), TNFSF13, transcription factor AP-1 (JUN), and IL-6 were associated with higher mRS score at 1 month after the onset of stroke. ITGA11, leucine-rich repeat neuronal protein 1 (LRRN1) and cell adhesion molecule-related/down-regulated by oncogenes (CDON) were negatively associated with mRS score (Table 4 and S8).

**Table 4:** Multivariable OR and 95% CI for the biomarkers associated with neurologic disability 1-month post-stroke according to modified Rankin Scale (mRS)

| **Biomarker** | **OR (95 CI%)** | **BH-adjusted P value** |
| --- | --- | --- |
| CCL23 | 3.08 (1.87-5.27) | <0.001 |
| CCL7 | 2.04 (1.38-3.05) | <0.001 |
| CDON | 0.45 (0.27-0.73) | 0.0477 |
| CKAP4 | 2.02 (1.33-3.10) | 0.0334 |
| CLEC4G | 2.38 (1.43-4.13) | 0.0334 |
| ENAH | 2.14 (1.45-3.21) | <0.001 |
| IL6 | 1.76 (1.33-2.37) | <0.001 |
| ITGA11 | 0.33 (0.18-0.61) | <0.001 |
| JUN | 1.9 (1.30-2.87) | 0.0334 |
| LRRN1 | 0.41 (0.23-0.72) | 0.0477 |
| LTBR | 2.02 (1.28-3.21) | 0.0477 |
| MATN2 | 2.45 (1.52-4.07) | <0.001 |
| PLAUR | 2.7 (1.63-4.60) | <0.001 |
| TNFSF13 | 1.98 (1.28-3.08) | 0.0477 |

Models adjusted for age and sex. Biomarkers names are available in Table S1.

### Association of inflammatory protein biomarkers with mortality at 1-month post-stroke

During 1 month of follow-up, 8 (8.89%) cases died. After correcting for FDR, no inflammatory proteins were found to be significantly associated with mortality (Table S9).

## DISCUSSION

To date, this is the first study to unravel novel inflammatory biomarkers in stroke patients in Africa using a proteomic discovery analysis. We have identified several candidate biomarkers that may underlie inflammatory processes as a response to cerebral ischemia, its severity, and neurologic disability during follow-up.

The majority of stroke-related differentially expressed biomarkers were linked to inflammatory response (e.g., TNFRSF11A, TNFSF11, CCL23 and IL-β). Among the upregulated proteins in stroke cases, AGRP was the most prominent. AgRP is best known as a hypothalamic neuropeptide that regulates whole-body metabolic adaptation to negative energy balance and catabolic states. Interestingly, experimental modulation of these neurons in mice has shown anti-inflammatory effects,^21,22^ including a distinct reduction in endotoxemia-induced circulating TNF-alpha.^22^ However, there is no substantial evidence that AgRP produced in the hypothalamus can be released into circulation. Nevertheless, AgRP has been detected in the bloodstream, with its mRNA and peptide observed in the adrenal medulla of both rodents and humans in glomus type I cells within the mouse carotid body, as well as in the testis and lung though confirmation of AgRP peptide production in these locations remains uncertain.^23–26^ Regardless of the source, circulating AgRP levels can be readily detected and have been reported to increase in rodents and humans in response to both obesity and prolonged fasting,^27–29^ conditions that share physiological stress responses, systemic inflammation, and compensatory anti-inflammatory mechanisms with post-stroke states. Nonetheless, the physio-pathological significance and the source of circulating AgRP remain unclear and will require future studies using preclinical models.

TNFRSF11A and its ligand TNFSF11 also known as Receptor activator of nuclear factor *κ*B (RANK) and (RANKL), respectively, are both expressed in activated microglia in the infarcted brain, promoting an inflammatory cascade by releasing several cytokines.^30,31^ One of these cytokines is colony-stimulating factor 1 receptor (CSF1R), a key regulator of microglial differentiation and survival.^32^ Notably, we show that CSF1R ligand, CSF1 is upregulated in ischemic stroke cases, aligning with other reports.^33^ Additionally, RANK is expressed in key regions of the brain associated with thermoregulation and is a key mediator of fever response to lipopolysaccharide-induced fever and pro-inflammatory cytokines, notably IL-1β and TNF-α.^34^ Elevated temperature following a stroke is associated with adverse outcomes, including the exacerbation of cerebral edema caused by the inflammatory cascade, increased intracranial pressure, and an increased likelihood of early mortality and poor functional outcomes.^35^ In line with other studies,^36^ we found that CCL23, a marker of immune cell trafficking, was associated with stroke severity and disability. Circulating CCL23 interacts with CC chemokine receptor 1 (CCR1), leading to the upregulation of several adhesion molecules that facilitate the migration of peripheral immune cells to the site of injury,^37^ which in turn contribute to cell damage.^38^ We have also found a strong association between C1qa and severity. C1qa is a subunit of the complement component 1q (C1q) molecule facilitating neurons’ phagocytosis by microglial cells promoting neurodegeneration.^39^

In contrast, stroke cases had lower plasma levels of integrin α-11, and it was inversely associated with stroke severity and disability. Integrins appear to have a role in the maintenance of blood-brain barrier integrity via interactions with extracellular matrix proteins and mediation of leukocyte adhesion to the endothelium of the blood-brain barrier.^40^ Most importantly, integrins regulate T regulatory (Treg) cells function,^41^ which contribute to the resolution and recovery of brain ischemia.^42^ However, the specific role of integrin *α*-11 in ischemia remains to be clarified. Although previous studies have shown that levels of IL-1β and IL-6 are upregulated post stroke,^43^ we observed lower levels of IL-1β in stroke cases compared to controls while IL-6 was found insignificantly different. Typically, IL-1β and IL-6 concentrations peak in the circulation in the first few days after stroke and then decline.^44,45^ Furthermore, expression of both cytokines is found to be elevated in the cerebrospinal fluid relative to peripheral circulation.^46^ Nevertheless, higher levels of IL-6 were associated with severity and disability after stroke.

There is a considerable interest in the contribution of specific toll-like receptors (TLR), which are activated by damage-associated molecular patterns, DAMPS (of extracellular, intracellular, and nuclear origin) in the ischemic cascade after stroke.^47^ In this context, previous clinical studies have found higher levels of TLR7 and TLR8 but not TLR3 or TLR9 in peripheral monocytes to be associated with a poor outcome following a stroke.^48^ We observed a higher TLR3 expression to be associated with a reduced likelihood of severe stroke. In experimental models, preconditioning with a TLR3 ligand prior to ischemic stroke has been demonstrated to decrease infarct volume and enhance neurological functions.^49^

In Africa, rapid urbanization, dietary changes and the burden of infectious diseases fabricate a pro-inflammatory phenotype along with genetic and environmental factors that contribute to disease progression and death.^6,50,51^ There is a disparity in studies investigating key factors of stroke outcomes and practices that may enhance the severity and prognosis of the disease, where standard care is absent.^51,52^ In this regard, finding key biomarkers of stroke outcomes might aid in patients stratification and developing new therapies.^53^ To the best of our knowledge, no large randomized controlled trials of anti-inflammatory agents have been conducted specifically for the prevention or improvement of stroke outcomes.^54^ Targeting IL-1R demonstrated promising improvement in stroke outcomes and reduction in inflammatory biomarkers when compared to placebo.^55^ It is important to note that these findings were derived from exploratory secondary analyses. Similarly, ApTOLL, a TLR4 antagonist, was associated with lower infarction volume, mortality and disability at 90 days compared to placebo.^56^ Other RCTs targeting inflammation in cardiovascular disease have reported outcomes related to stroke. For instance, colchicine decreased the risk of incident stroke in people with coronary artery disease.^57^ While canakinumab failed to reduce stroke events, probably due to lower number of events,^58^ evidence from experimental stroke models suggests that targeting IL-1β by canakinumab might improve stroke outcomes.^59^

### Limitations

Several limitations should be acknowledged in the current study. Firstly, the observational nature of this study limits our ability to establish causal relationships. The samples for measuring the plasma inflammatory protein profile were collected at admission among acute ischemic stroke cases with a duration of onset of stroke symptoms varying between 1 and 8 days. This variability may reflect temporal and differential variations in the timing of sample collection. The Olink panel utilized in this study primarily consisted of biomarkers associated with inflammation and we cannot describe association with non-inflammatory biomarkers. We did not identify any associated proteins with infarct volume or mortality, which may be attributed to the limited sample size in this study. Additionally, the restricted sample size precluded any stratification analysis. Finally, the proteomics assay in this study lacks standardized concentration units, rendering comparisons with clinically established cutoffs challenging.

### Conclusion

Biomarkers of inflammation are associated with ischemic stroke severity and disability in indigenous Africans. Their potential to predict stroke outcomes or serve as therapeutic targets in this population warrants further investigation.

## Data Availability

The data for this study can be accessed upon request from the corresponding author.

## Authors Contributions

Prof Owolabi and Mr Morsi had full access to all of the data in the study and takes responsibility for the integrity of the data and the accuracy of the data analysis.

*Concept and design:* Morsy, Sarfo, Maffia, Owolabi.

*Acquisition, analysis, or interpretation of data:* Morsy.

*Drafting of the manuscript:* Morsy, Sarfo, Maffia, Owolabi.

*Critical review of the manuscript for important intellectual content:* All authors.

*Statistical analysis:* Morsy.

*Obtained funding:* Maffia.

Administrative, technical, or material support: The SIREN team.

*Supervision:* Maffia, Owolabi.

## Supplemental Material

STROBE Checklist

Figure S1

Tables S1-S9

## Nonstandard Abbreviations and Acronyms

AgRP: Agouti-related peptide
CCL23: Chemokine (C-C motif) ligand 23
IL: Interleukin
ITGA11: Integrin α-11
mRS: Modified Rankin Scale
NIHSS: National Institutes of Health Stroke Scale
RANK: Receptor activator of nuclear factor *κ*B
RANKL: Receptor activator of nuclear factor *κ*B ligand
SLS: Stroke Levity Scale
TLR: Toll-like receptor
TOAST: Trial of ORG 10172 in Acute Stroke Treatment

